# Evaluation of saliva self-collection devices for SARS-CoV-2 diagnostics

**DOI:** 10.1101/2021.02.01.21250946

**Authors:** Orchid M. Allicock, Mary E. Petrone, Devyn Yolda-Carr, Mallery Breban, Hannah Walsh, Anne E. Watkins, Jessica E. Rothman, Shelli F. Farhadian, Nathan D. Grubaugh, Anne L. Wyllie

**Affiliations:** Department of Epidemiology of Microbial Diseases, Yale School of Public Health, New Haven, CT 06510, USA; Department of Medicine, Section of Infectious Diseases, Yale School of Medicine, New Haven, CT, 06510, USA

## Abstract

There is an urgent need to expand testing for SARS-CoV-2 and other respiratory pathogens as the global community struggles to control the COVID-19 pandemic. Current diagnostic methods can be affected by supply chain bottlenecks and require the assistance of medical professionals, impeding the implementation of large-scale testing. Self-collection of saliva may solve these problems, as it can be completed without specialized training and uses generic materials. In this study, we observed thirty individuals who self-collected saliva using four different collection devices and analyzed their feedback. Two of these devices, a funnel and bulb pipette, were used to evaluate at-home saliva collection by 60 individuals. All devices enabled the safe, unsupervised self-collection of saliva. The quantity and quality of the samples received were acceptable for SARS-CoV-2 diagnostic testing, as determined by RNase P detection. Here, we demonstrate inexpensive, generic, buffer free collection devices suitable for unsupervised and home saliva self-collection.

## Introduction

Over a year since COVID-19 was declared a pandemic, the demand for testing remains high. Even with the rollout of several vaccines, successful control strategies still depend upon the availability of reliable, scalable testing programs. Self-collection of saliva for SARS-CoV-2 testing can facilitate these. Numerous studies have shown that saliva is an equally sensitive substrate for the detection of SARS-CoV-2 RNA as nasopharyngeal swabs (Hanson et al., 2020; Tan et al., 2021; Vogels et al., 2020; Wong et al., 2020; Wyllie et al., 2020). Unlike sampling with nasopharyngeal swabs, self-collection of saliva is non-invasive and does not require specialized training to perform (Marty et al., 2020). Moreover, SARS-CoV-2 RNA is stable in saliva at a broad range of temperatures and for an extended period of time, obviating the need for cold chain storage and preservatives or buffers that increase the costs of collection (Ott et al., 2021).

While saliva has been used as a diagnostic testing substrate for pathogenic antibodies (Drobnik et al., 2011; Korhonen et al., 2014; Reynolds and Muwonga, 2004), its utility in viral pathogen detection has been limited to viruses like human immunodeficiency virus (Yapijakis et al., 2006), measles, mumps, and rubella (Jin et al., 2002), human papillomavirus (Adamopoulou et al., 2008), Epstein-Barr virus (Idesawa et al., 2004) and certain viral co-infections (Kim et al., 2017; Robinson et al., 2008; Yoon et al., 2017), all strictly in research settings. Before 2020, the only PCR-based diagnostic test using saliva (saliva swabs) approved or authorized by the FDA was for the detection of human cytomegalovirus in babies (FDA, 2018). Through the development of saliva-based diagnostic tests, COVID-19 testing became more accessible.

Despite its advantages, if saliva is collected improperly, it is difficult to handle in the laboratory (Landry et al., 2020). Improper self-collection may also pose a safety risk if potentially biohazardous materials are mishandled. Therefore, it is essential that self-collection of saliva is safe and can produce testable samples. Equally important is establishing the acceptability of self-collection among the general public because methods that are deemed uncomfortable, difficult, or confusing are unlikely to gain traction in the population.

In this study, we evaluated the experience of thirty individuals who self-collected saliva using four different saliva collection devices: a P1000 pipette tip, a Salimetrics Saliva Collection Aid (Salimetrics LLC, Pennsylvania, USA), a funnel, and a bulb pipette (**Figure 1a**). We found that all four devices enabled the consistent and safe collection of true saliva that was acceptable for SARS-CoV-2 diagnostic testing with a RT-qPCR-based assay (Vogels et al., 2020). Using this information we next evaluated the suitability of both a funnel and a bulb pipette for unsupervised at-home saliva collection. Our findings demonstrate the suitability of multiple device options for use in saliva self-collection kits. This variety not only helps to avoid supply chain bottlenecks but could also promote broader acceptance of this method by improving the ease of self-collection and of sample processing in the laboratory.

**Figure 1:**
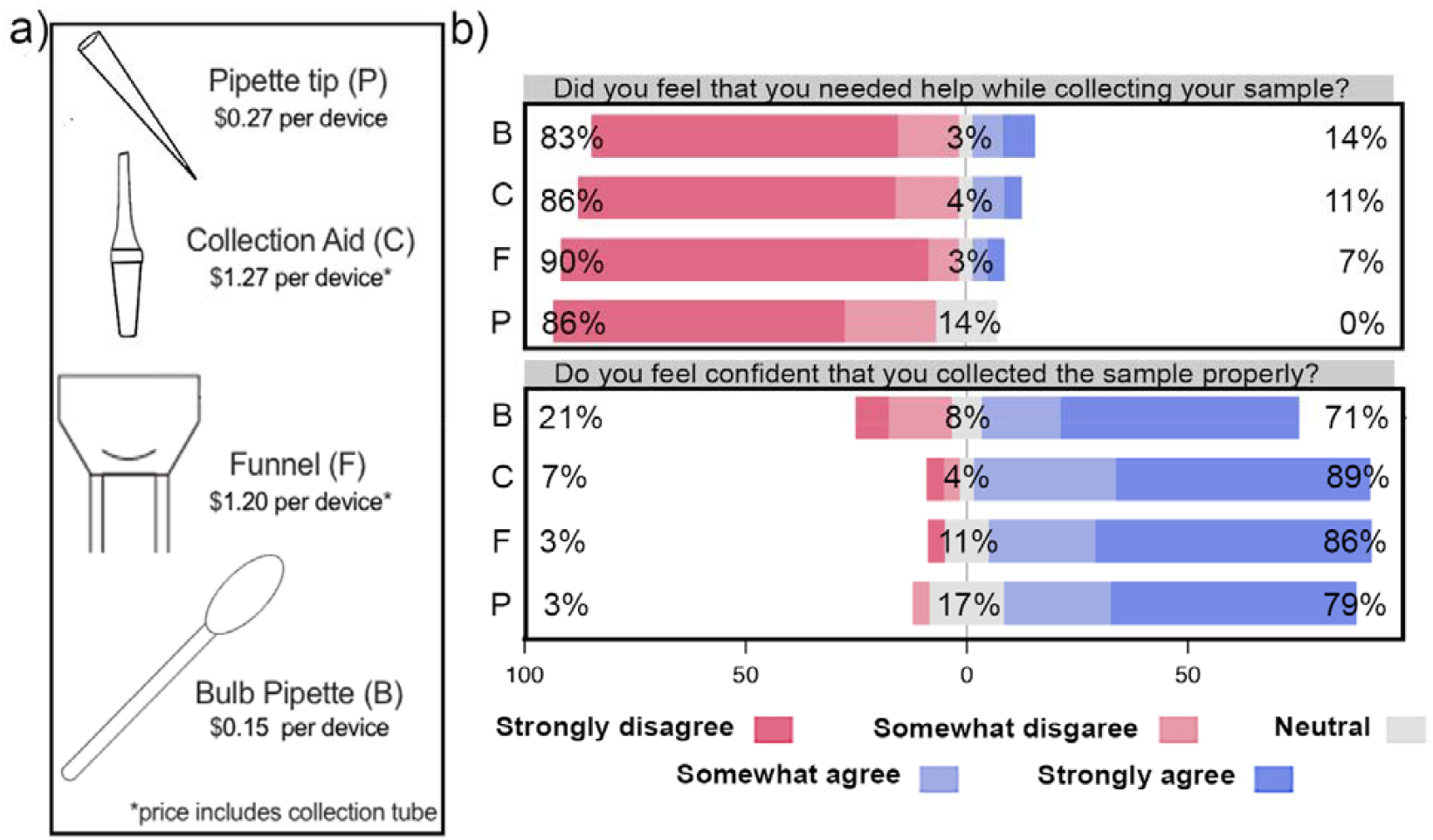
Collection devices are inexpensive, easy to use, and yield testable samples. Survey responses were reported from strongly disagree to strongly agree. (**a**) The four collection devices tested are inexpensive and provide users with a range of features to choose from. Prices at time of publication are shown in US dollars. (**b**) Participants reported being self-sufficient and confident in their ability to correctly collect saliva samples (from **Figure S2**). The questions are displayed above the corresponding graphs. The percentage response value for each device is shown above each bar. Two sets of participant responses were excluded because one participant did not provide a response for all four devices and one did not understand the response scale. *Abbreviations: P = pipette tip, C = collection aid, F = funnel, B = bulb pipette*.

## RESULTS

### All four saliva collections devices were deemed usable by the study participants, but individual preference influenced their relative acceptability

We aimed to enroll participants who represented a range of racial and educational backgrounds (**Table 1**). In 100% of the observed collections, study participants appeared confident in their ability to complete the collection correctly (**Figure S2**). The majority of participants (93%) understood the importance of following the instructions carefully to avoid incorrect test results, and during only two collections (1.67%), participants appeared to not adequately follow these instructions for proper sample collection (**Figure S2b**).

**Table 1.** **Demographic characteristics of the participants from the pilot study and the at-home saliva kit study.**

| Category | Pilot study (n=30)<br>n (%) | At-home kit (n=60) |  |  |
| --- | --- | --- | --- | --- |
|  |  | Overall (%) | Bulb (%, n= 31) | Funnel (%, n=29) |
| **Sex** |  |  |  |  |
| Male | 11 (37) | 28 (47) | 14 (45) | 14 (48) |
| Female | 19 (63) | 32 (53) | 17 (55) | 15 (52) |
| **Age** |  |  |  |  |
| 18-29 | 7 (23) | 13 (22) | 7 (23) | 6 (21) |
| 30-39 | 16 (53) | 27 (45) | 11 (35) | 16 (16) |
| 40-49 | 4 (13) | 5 (8) | 4 (13) | 1 (3) |
| 50-59 | 0 (0) | 5 (8) | 3 (10) | 2 (7) |
| 60-69 | 1 (3) | 6 (10) | 4 (13) | 2 (7) |
| 70+ | 2 (7) | 4 (7) | 2 (6) | 2 (7) |
| **Education** |  |  |  |  |
| High School/GED | 2 (7) | 15 (25) | 9 (29) | 6 (21) |
| Bachelors | 7 (23) | 21 (35) | 11 (35) | 10 (34) |
| *Masters* | 10 (33) | 11 (18) | 7 (23) | 4 (14) |
| *PhD/MD* | 11 (37) | 13 (22) | 4 (13) | 9 (31) |
| **Race** |  |  |  |  |
| *Black/African American* | 4 (13) | 10 (17) | 6 (19) | 4 (14) |
| *Hispanic/Latino* | 4 (13) | 10 (17) | 5 (16) | 5 (17) |
| *Asian/South Asian* | 6 (20) | 7 (12) | 4 (13) | 3 (10) |
| *White* | 15 (50) | 33 (55) | 16 (52) | 17 (59) |
| *Native American* | 1 (3) | 0 (0) | 0 | 0 |

Of the 10 participant survey questions, only Question 5 (“Was collecting the sample difficult in general?”) varied statistically significantly across devices; however, this question was found to not be internally reliable (**Table S1, S2**). In this case, the bulb pipette scored the least favorably (mean = 3.1) compared to the other devices (pipette tip, mean = 2; funnel, mean = 2.3; collection aid, mean = 1.7) (**Figure S2**). Participants commented that the bulb pipette introduced bubbles and caused discomfort if it suctioned the inside of their mouth (**Table 2**). Despite this feedback, all participants provided a sufficient volume of saliva for testing with all four devices, the majority did not think they required assistance during the sample collection (93%), and in only 18 collections (16%), participants did not feel confident that they had collected the sample correctly with the bulb pipette (**Figure 1**). Similarly, observers reported that the majority of participants did not appear to struggle with the collection process (115/120, 95.8%, **Figure S2b**).

**Table 2.**
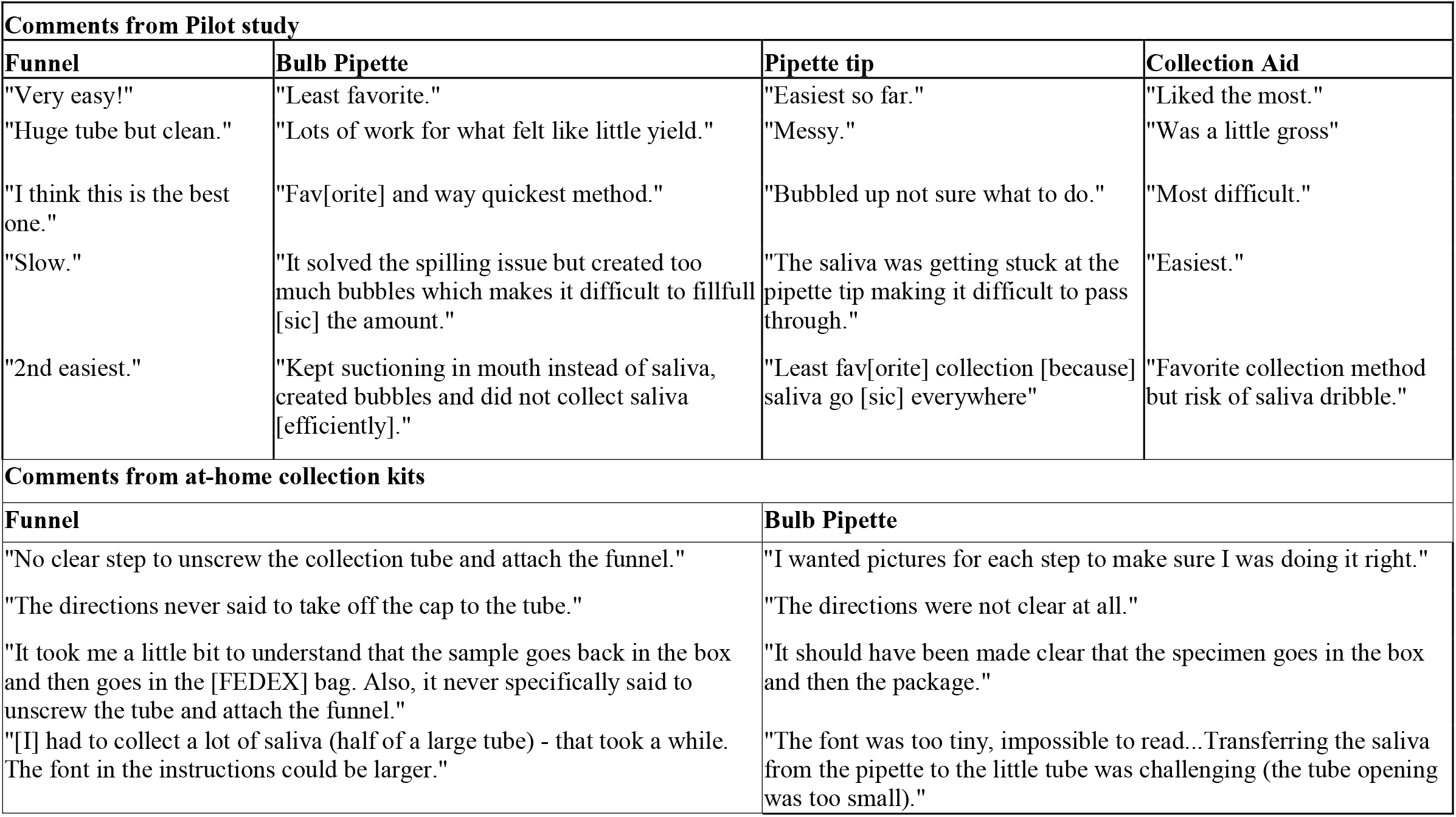
Participant observations and experiences with the unobserved collection. Survey comments from participants from pilot study and the at-home collection kits. The comments presented here are representative but not exhaustive. *Abbreviations:* No. = number.

In addition to answering the survey questions, participants were given the opportunity to provide general feedback. Each device received a range of comments from participants reflecting differences in personal preference (**Table 2**). For example, though the bulb pipette received the largest number of negative comments (n=11), one participant stated it was their favorite of the four devices. Interestingly, there was no general consensus around an overall preferred device; however, the size of the devices was a common theme among participant feedback. Some participants (4/30, 13%) found the pipette tip and collection aid to be too small, whereas the large size of the funnel and its collection tube were noted to be an advantage. More research is needed to determine which types of devices may be most suitable for specific demographic groups, but it is likely that providing a range of options will promote the general acceptability of saliva self-collection for pathogen diagnostic testing.

### Unsupervised saliva collection can be reliably conducted at home

In order to achieve diversity in the demographics of the participants, we selected 84 of the 246 participants who consented to unsupervised at-home saliva collection study, based on age, sex, race and educational status. The participants were sent self-collection kits containing either a funnel (n=43) or bulb transfer pipette (n=41) to aid saliva collection. Of those distributed, 66 kits were returned, however 6 participants did not complete the survey, so were excluded from the study. Overall, survey responses following unsupervised collection were favorable (**Figure 2b, Figure S4**). Participants reported feeling confident with carrying out self-collection properly and that the process was not difficult. Importantly, study participants clearly understood the required process of sample collection, with 100% of participants acknowledging that they understood not to eat/drink/smoke prior to collecting the sample, and 88.33% understood that incorrect sampling could result in false results (**Figure S4**). There were slight differences in the user experience between bulb pipette kits and the funnel kits; 16% of the participants found that the sample collection was difficult with the bulb pipette as compared to only 7% of the participants using the funnel.

**Figure 2.**
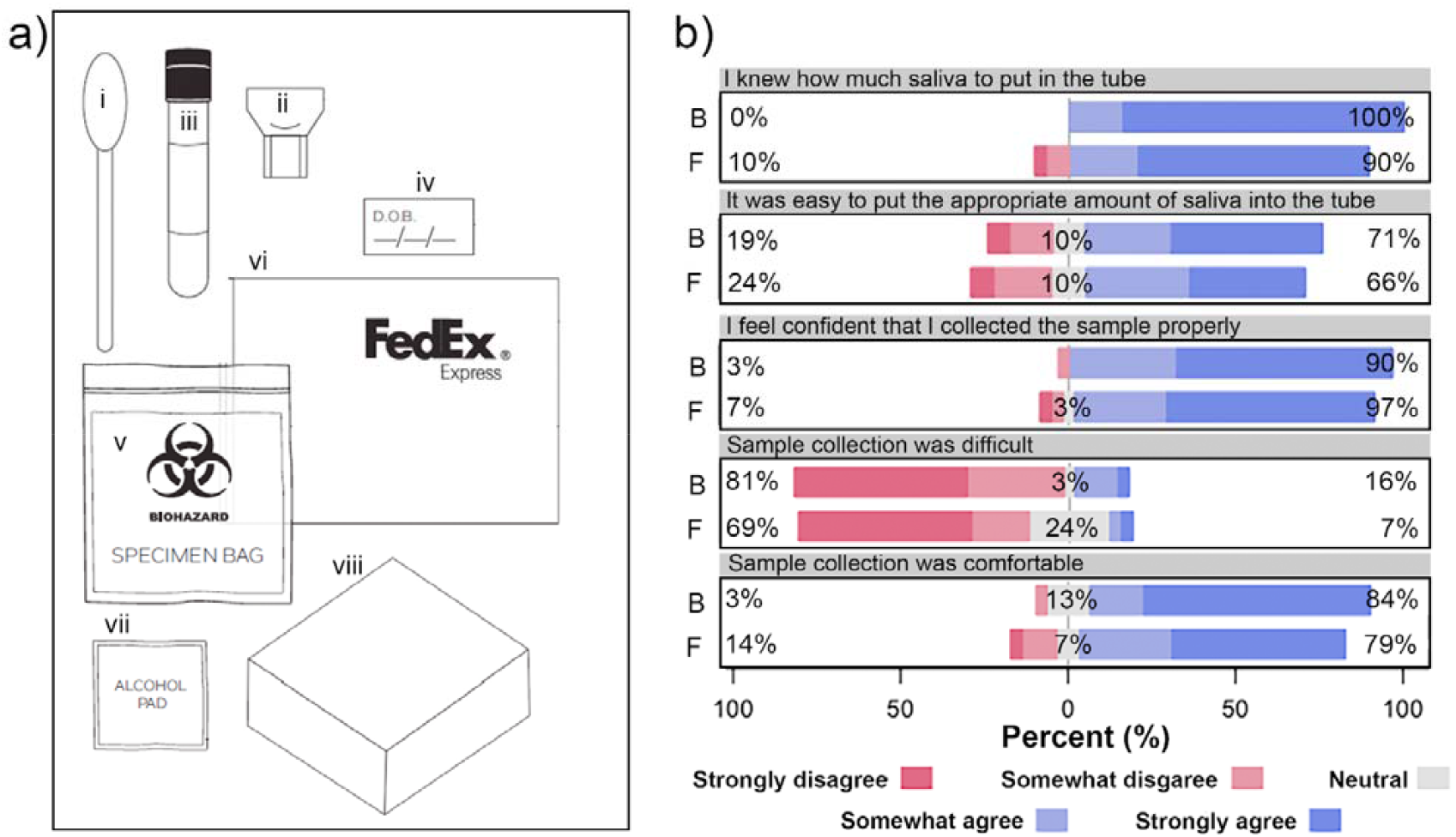
At-home saliva collection kit components suitable for sample collection. **a)** Each of the participants were sent an at-home collection kit comprised of either a funnel (i) or bulb pipette (ii) with a labeled screw-cap tube (iii), patient identifier sticker (iv), biohazard collection bag with absorbent sheet (v), FedEx UN 3373 Pak (vi), an alcohol pad (vii), and box for return shipment (viii). **b)** Participant confidence in at-home self-collection of saliva when using either a funnel or bulb pipette (from **Figure S4**). Survey responses were reported on a scale of 1 (strongly disagree) to 5 (strongly agree). Overall, there was no significant difference between the collection devices in relation to the participant’s confidence and ability to use either device. The questions are displayed above the corresponding graphs. *Abbreviations: F = funnel, B = bulb pipette*.

### Self-collection of saliva was safe and yielded testable samples

Ensuring the proper handling of potentially biohazardous material is an essential consideration for saliva self-collection to be implemented on a large scale. Specifically, contamination of the collection tube with virus-infected saliva poses the greatest health and safety risk for this method.

Some participants did contaminate the outside of their collection tubes with saliva during the pilot collection (27.8%) and the at-home kit study (21.7%), but participants from the pilot study were observed sanitizing the collection tube with an alcohol wipe in accordance with the provided instructions and the majority of at-home study participants reported understanding what to do in this situation. Additionally, as directed in the written instructions, 87% of participants in the pilot study washed or sanitized their hands before and after completing the collections. Regardless, strict sample handling safety precautions should be applied by all testing laboratories when receiving any clinical sample type.

Our secondary objective was to compare the quality of samples collected using each device. We found that all of the samples received (both unobserved as well as unsupervised at-home self-collection) were of sufficient quality for testing with SalivaDirect (Vogels et al., 2020), demonstrating how true saliva, which naturally pools in the mouth, can be easily handled in the laboratory. Specifically, laboratory survey responses confirmed that 100% of the samples collected during the pilot study were easy to pipette and of sufficient volume (>0.5 mL) (**Figure 3, Figure S3**). Slight discoloration was noted in 18 samples (15%) and food particles were observed in 20 samples (5 participants, 16.7%), but these did not affect test results. No sample tested positive for SARS-CoV-2. The average cycle threshold (Ct) value for the negative control, RNAse P (RP), was within the expected range (23-28 Cts) (Wyllie et al., 2020) for the majority of samples from the pilot study (73%), indicating that the use of different collection methods did not interfere with the diagnostic assay (**Figure 3**). We did not find a significant difference between matched samples across devices using one-way ANOVA (**Figure. S3**).

**Figure 3.**
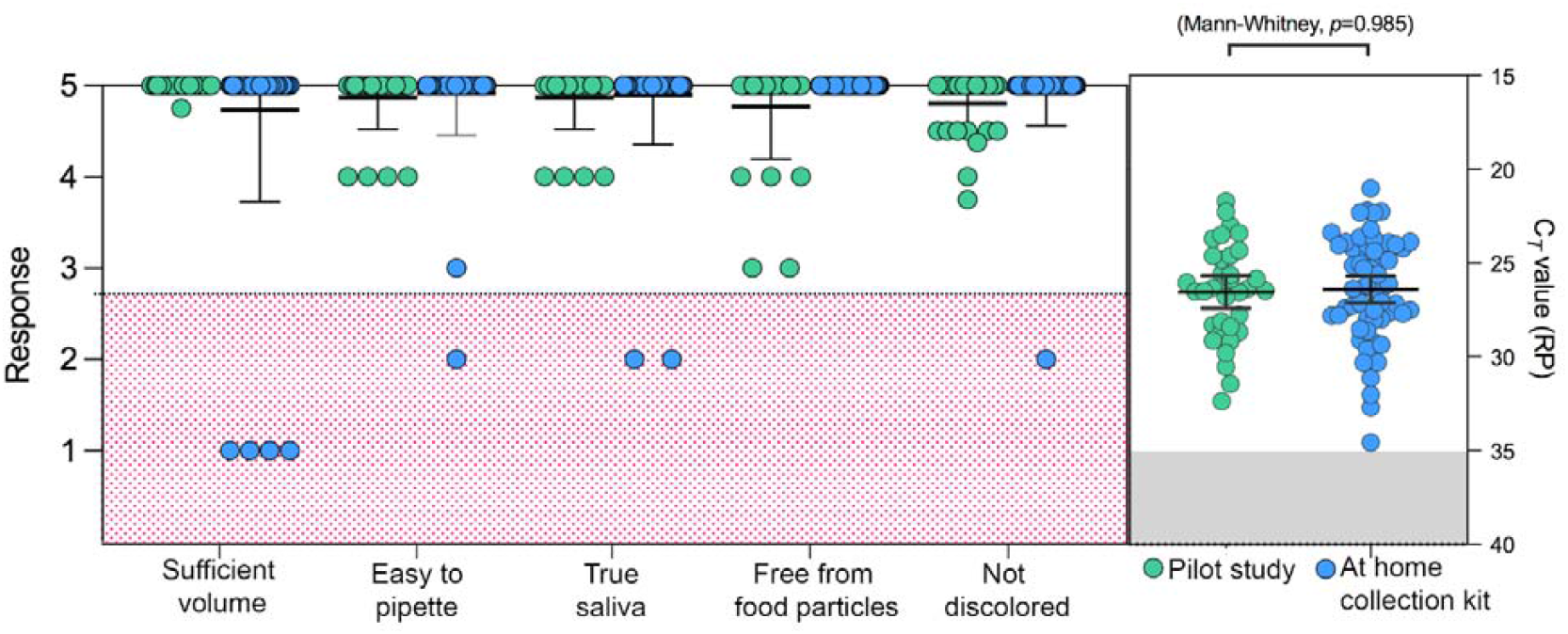
The quality of the samples was adequate for testing with a PCR-based assay. Laboratory survey questions pertaining to the quality of the samples are shown on the x-axis (from **Figures S3, S5**). Data points represent the mean response, green dots represent samples collected from the pilot study and blue dots represent samples collected from the at-home collection kit. Survey responses were reported on a scale of 1 (strongly disagree) to 5 (strongly agree). Samples with less favorable responses are highlighted in red. Mean and standard deviation (st. dev.) are shown in black. The graph on the right shows the cycle threshold (Ct) values for the internal control RNAse P (RP) from each of the saliva samples submitted. The blue and green dots represent Ct value per participant. Ct values over 35 are considered invalid and is highlighted in gray. P-value is shown using one-way Mann-Whitney. Mean and standard deviation (st. dev.) are shown in black.

The overall quality of the saliva samples from the at-home kit study was also acceptable, but with slight differences between the two collection devices. Of the 60 samples returned, 6.7% (n=4) contained less than 0.5 mL saliva (half of the 1 mL tube provided), all from participants given the bulb pipette collection kit (**Figure 3, Figure S4**). Despite this, all 4 samples were sufficient for testing, containing 100-450 µL of saliva. Besides low volume, the quality of the samples collected with the bulb pipette was high, with 100% of samples easy to pipet, free from food particles, not discolored and consisting of only true saliva. On the other hand, while 100% of the samples returned from the participants with the funnel collection kit were of sufficient volume, 2/29 samples might not have been “true saliva” and as a result were difficult to pipet. In addition, one of the samples was slightly discolored.

## Discussion

To combat the ongoing outbreaks of SARS-CoV-2, mass testing strategies which are cost-effective and free from supply chain disruptions are essential. Additional major barriers to frequent testing result from a need to schedule appointments at facilities staffed with trained personnel or testing aversion to swab-based methods. Scaling up the use of saliva self-collection as a routine diagnostic tool can expand access to testing for SARS-CoV-2 and could be reliably performed in workplaces, schools or college dormitories where regular testing is essential for safe day-to-day operations. To support these efforts, we aimed to identify saliva collection solutions with generic components without sacrificing the comfort of the participants or the effectiveness of collection. Results from this study demonstrate the usability and efficacy of several simple saliva collection methods for SARS-CoV-2 detection. Importantly, all of the devices promoted the collection of “true” saliva, which was acceptable for handling in the laboratory, and were deemed usable by our participants.

The data collected from the pilot study was used to inform our selection of the bulb pipette and funnel as the saliva collection devices for the at-home saliva collection kits. Though there was no clear preference in devices based on demographic factors like sex, education level, ethnicity or age, some of the older participants had issues with saliva collection using the bulb pipette. More studies can be done to specifically assess the usability of the collection devices in these specific populations. The availability of the option for unsupervised sample collection for COVID-19 testing could result in up to one-third more symptomatic persons seeking testing, especially in those populations of individuals who are at high risk for contracting the infection, or those who are unable/unwilling to go into clinical settings (Siegler et al., 2020). With more options available, individuals can select kits according to their needs and limitations.

We did not directly compare the self-collection process with the aid of a collection device to the process without a device, but the ability to collect true saliva in simple wide mouth tubes has been previously demonstrated (Byrne et al., 2020; Wyllie et al., 2020). Wide-mouth tubes are not conducive for large-scale testing in labs with limited space or when sample processing requires the use of a liquid-handling robot, a piece of equipment present in most large clinical laboratories. Therefore, the collection devices we tested allow for an easy collection process into smaller tubes that are likely more amenable to the majority of laboratory procedures. Importantly, results from our study also demonstrate that these devices do not inhibit RNA-extraction free, RT-qPCR based diagnostic assays.

This study also evaluated the instructions for reliable saliva self-collection. The majority of the participants had no additional feedback, and the few comments we did receive were all related to the kit instructions, involving font size, mailing instructions and device assembly (see table 3). This slight confusion was reflected in the participant survey responses, where 35% of participants were unsure of what to do if saliva came into contact with the outside of the tube and 26% were unsure of what to do if they had any questions. This feedback highlighted the need to further refine the instructions in order to decrease the likelihood of errors in saliva collection and improve the sample collection experience. Additionally, visual materials such as a video outlining the sample collection and shipping process could be helpful in future iterations of the kits.

Even with ongoing vaccination campaigns, widespread, routine testing for SARS-CoV-2 will remain a staple of public health disease control strategies for at least another year. For this, unsupervised saliva collection permits feasible, scalable, and affordable testing solutions.

## Limitations of the study

While the sample size of the pilot study was small, and a majority of study participants held a college degree or higher, similar results were obtained when we enrolled a larger, more demographically diverse cohort for the unsupervised, at-home evaluation. It is important to note that we did not enroll individuals under the age of 18 and therefore cannot draw conclusions around the usability of these devices in children. However, large-scale pathogen surveillance testing involving self-collected saliva samples from school-aged children have been executed for SARS-CoV-2 and other pathogens (*Streptococcus pneumoniae)* (Bi et al., 2021; Wyllie et al., 2014).

Overall, the response to the collection devices were favorable. However the sample size was too small to determine if there are age-specific preferences in collection devices. More studies can be done to assess the utility of different collection devices in select populations.

## Data Availability

De-identified survey responses and source data are available at doi:10.17632/x2mv2ctm7c.1 and in the supplement.

## Author contributions

A.L.W. and N.D.G. conceived the study. H.W., M.E.P. and S.F.F. assisted with the coordination and execution of the study. M.E.P, O.M.A., D.Y-C., and M.B. observed the collections. M.B., D.Y-C., O.M.A. and A.E.W. performed the diagnostic tests. M.E.P., O.A. and D.Y-C. analyzed the data. J.E.R. assisted with the design of the statistical analysis. M.E.P., O.A., D.Y-C., N.D.G., and A.L.W. wrote and edited the manuscript.

## Acknowledgements

We thank the study participants for their time and cooperation. We also thank Una Pipic, Jessica Metti, Monisha Appalarju and the team at Tempus for their support. This work was funded by Tempus Labs, Inc (N.D.G and A.L.W), Yale Center for Clinical Investigation TL1 TR001864 (M.E.P.) and Fast Grant from Emergent Ventures at the Mercatus Center at George Mason University (N.D.G and A.L.W).

## Methods

### Ethics

This study was conducted in accordance with an Institutional Review Board protocol reviewed and approved by the Yale University Human Research Protection Program (IRB Protocol ID: 2000028394).

### Study design

For the initial evaluation of unobserved saliva collection, thirty participants between the ages of 20 and 80 years were enrolled. Individuals who had previously provided a saliva sample, who had relevant, career-level laboratory experience, or who were experiencing symptoms of respiratory infection were excluded from enrollment. Once informed consent was provided, participants received a collection kit containing (1) the four saliva collection devices (**Figure 1a**), (2) corresponding collection instructions, (3) a biohazard bag, and (4) five alcohol wipes. Participants self-collected four saliva samples consecutively and in a randomized order. Members of the study team observed these collections via a video platform with minimal interaction with the study participant. The observer turned off video and audio on their device for the duration of the four collections and provided no instructions on sample collection. Following each collection, both the observer and the study participant completed a survey about the experience, scoring responses on a scale of 1 (strongly disagree) to 5 (strongly agree) (**Figure S1**).

An additional 60 participants were recruited into the study through an online, social media post to evaluate unsupervised at-home saliva collection. Participants were required to be at least 18 years of age, reside in the contiguous United States with no previous experience with providing saliva for diagnostic testing. Participants provided demographic data and were consented via an online form to limit direct contact with study participants, and to replicate an unsupervised at-home collection as closely as possible. Study participants were selected from consenting individuals to ensure a diverse range of age and race. Study participants were mailed an at-home self-collection kit containing a saliva collection device, a collection tube, collection instructions, a biohazard bag, an alcohol wipe and a FedEx envelope for sample return (**Figure 2a**). Samples returned to the laboratory were stored at 4°C for up to 4 days until testing.

### Sample testing

All saliva samples (n = 183) were tested for a region of the SARS-CoV-2 nucleocapsid gene (N1) and human RNase P (RP) using the SalivaDirect protocol (Vogels et al.). A laboratory survey assessing the quality of each sample was completed by the technician during testing.

### Statistical analysis

Participant, observer, and laboratory survey questions were tested for internal reliability with Cronbach’s alpha using R v.4.0.2. Significant statistical differences across the 4 devices were calculated using one-way ANOVA in GraphPad v.8.4.3. Participants who did not provide a response for all four devices were excluded from the analysis for the corresponding question (maximum of 6 for question 10). For the laboratory surveys, responses to questions 2, 3, and 4 were identical across devices and therefore could not be assessed using one-way ANOVA. For the at-home self-collection of saliva, the differences between the bulb pipette and funnel kits were assessed using the Mann-Whitney test in GraphPad v.9.1.0.

## Supplementary Figures

**Figure S1.**
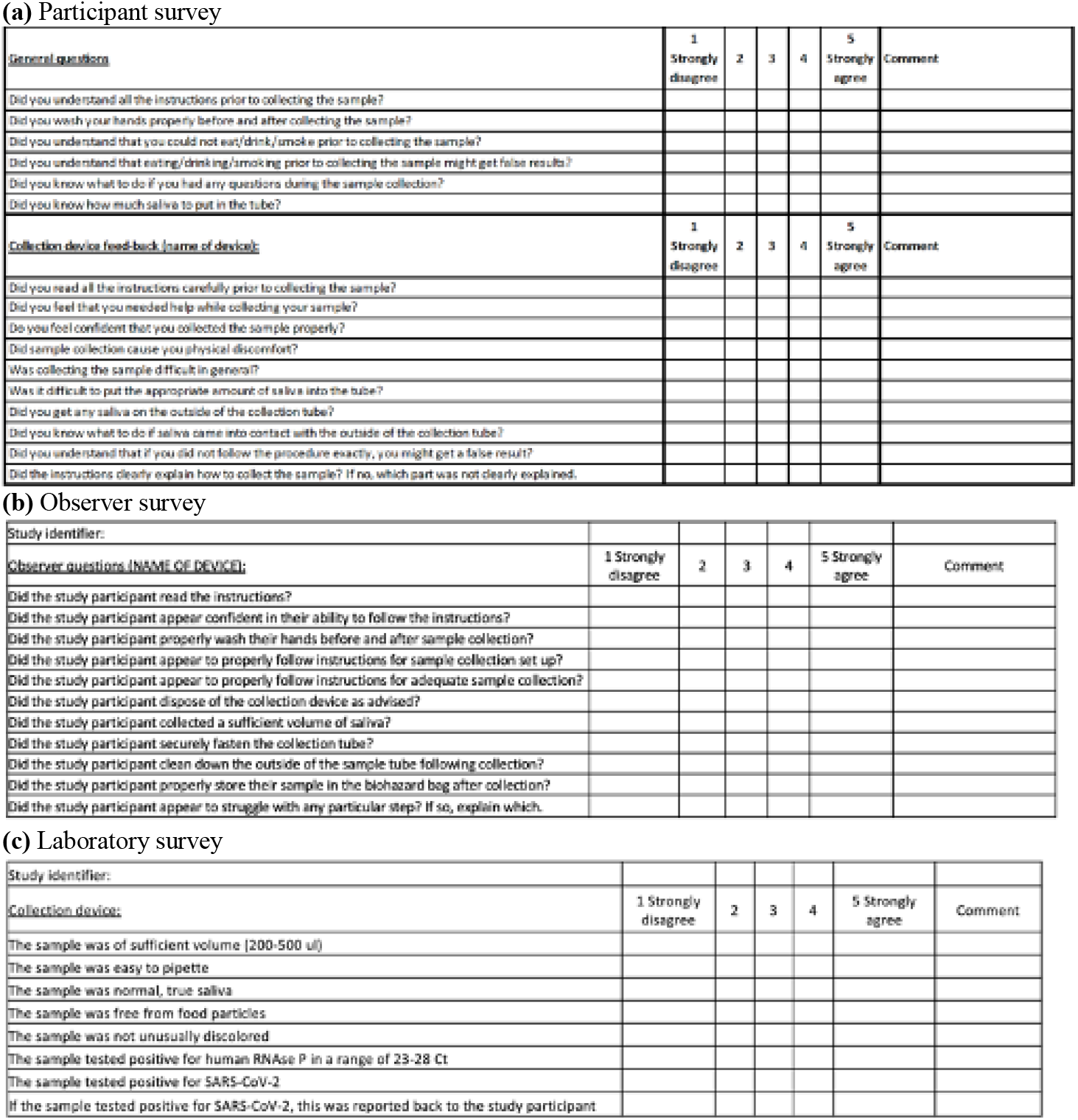
Survey questions posed to participants, observers, and laboratory personnel. For the pilot study, parts a), b) and c) were used. For the at-home kit, only a) and c) were used. Responses were given on a scale of 1 (strongly disagree) to 5 (strongly agree).

**Figure S2.**
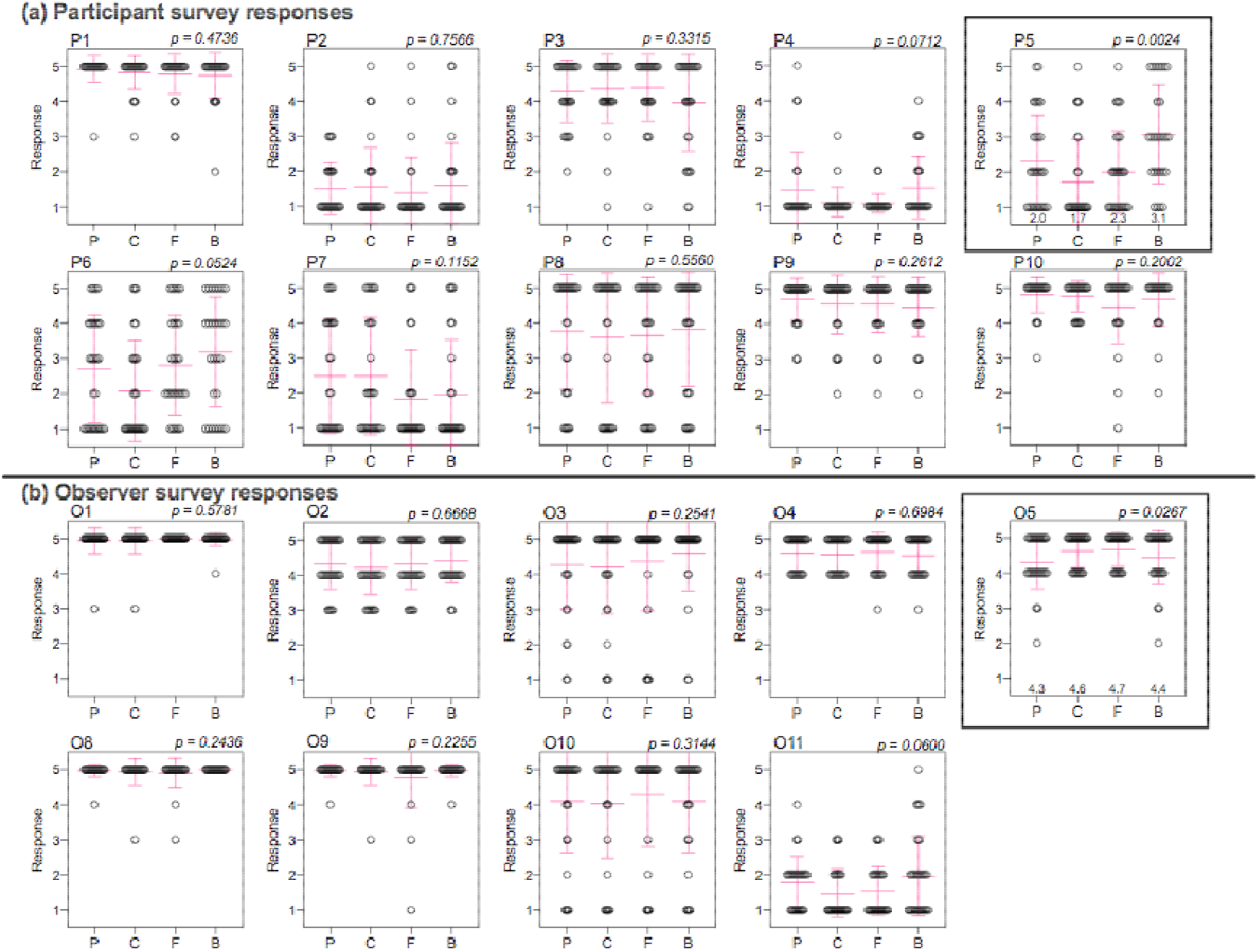
Responses to participant and observer surveys (related to Figure 1). Mean and standard deviation are marked in pink. Survey data were analyzed using one-way ANOVA. Responses to two questions (P5 and O5) differed significantly across devices and are denoted with black boxes. The numbers shown on the x-axis of those graphs are the mean response value. *Abbreviations: P = pipette tip, C = collection aid, F = funnel, B = bulb pipette*.

**Figure S3.**
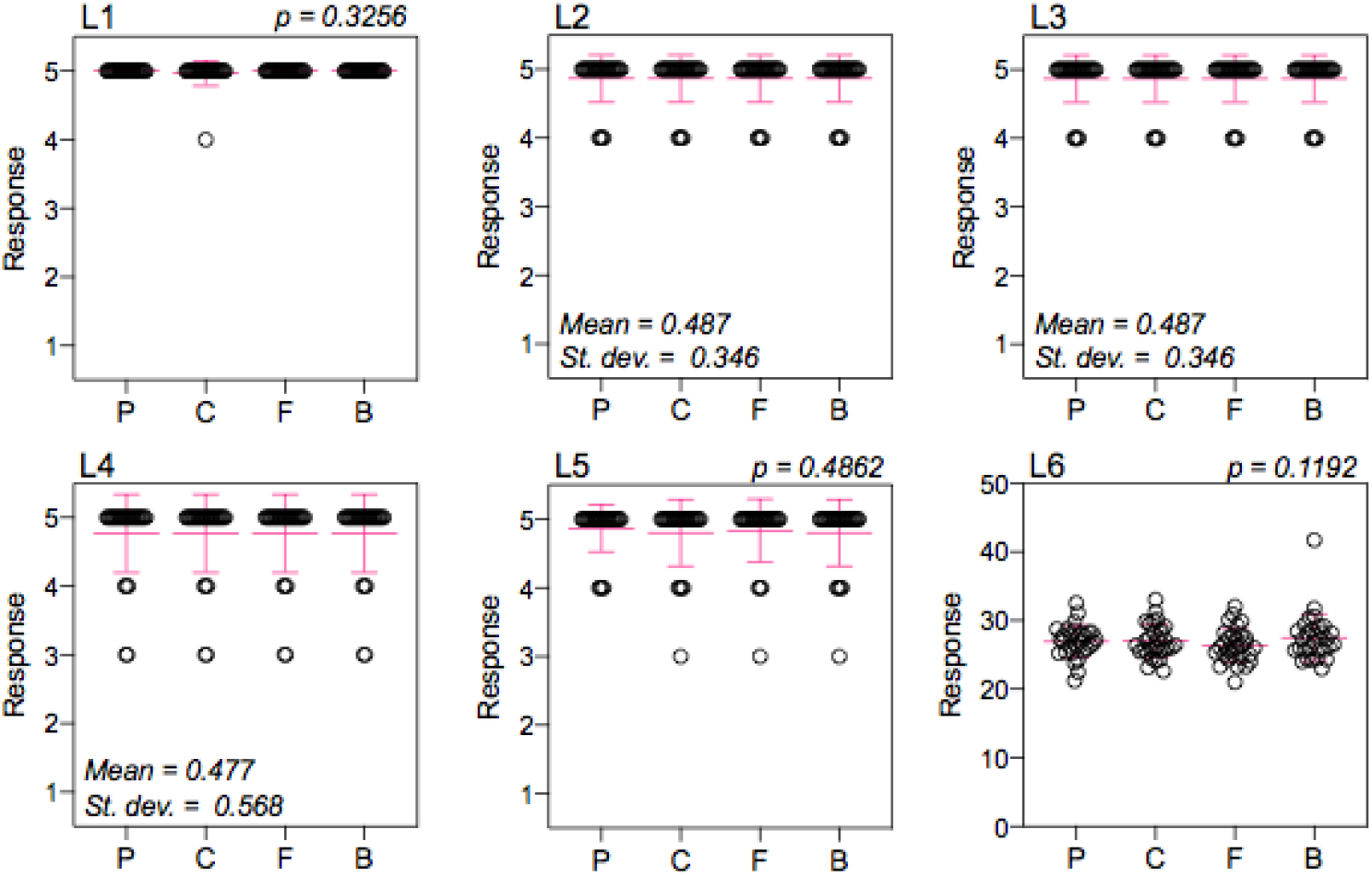
Responses to laboratory survey. P-values are shown for questions that could be assessed using one-way ANOVA. Mean and standard deviation (st. dev.) are shown for questions where responses were identical across devices. *Abbreviations: P = pipette tip, C = collection aid, F = funnel, B = bulb pipette*.

**Figure S4.**
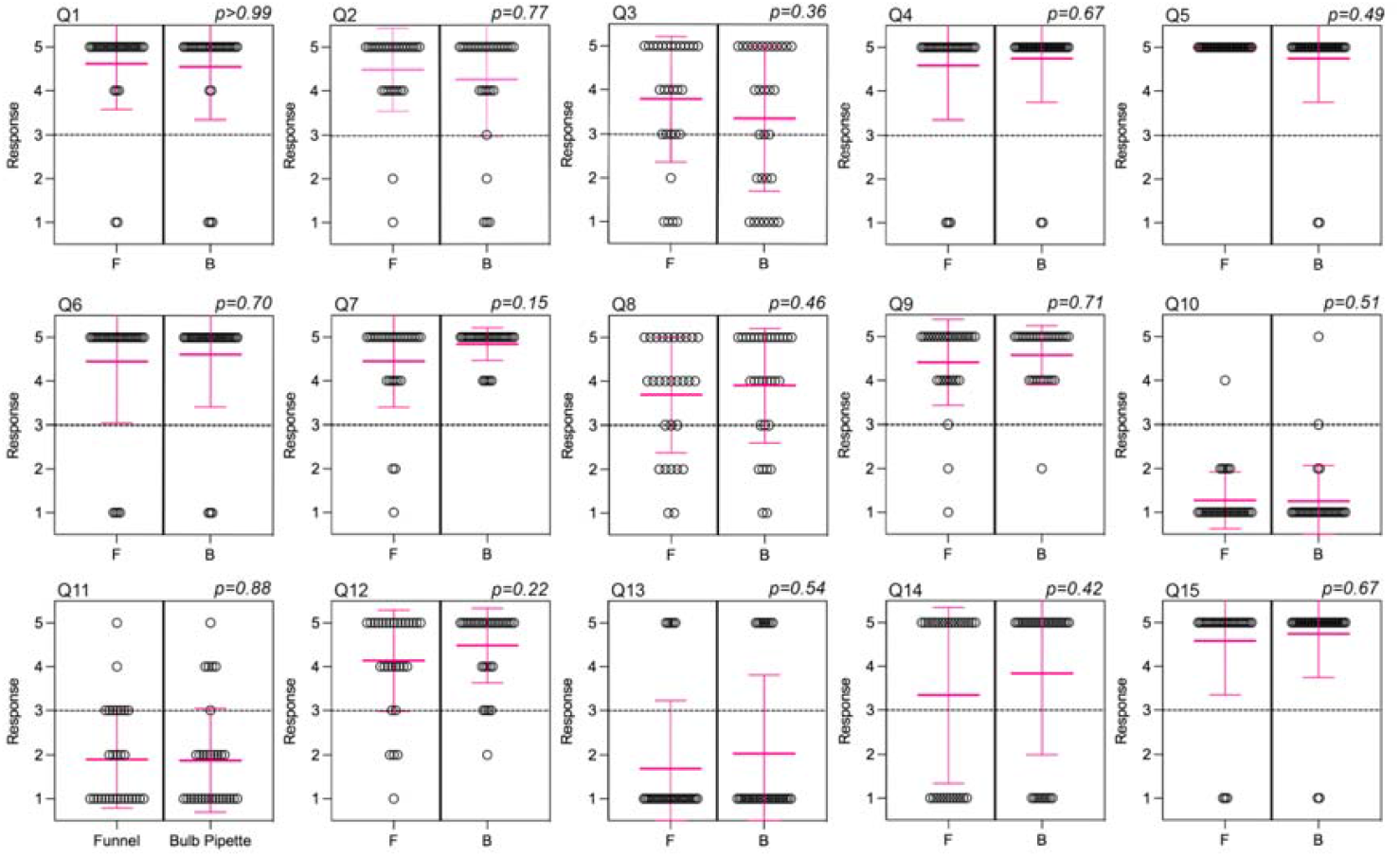
Responses to participant surveys for at home collection kit. Mean and standard deviation are marked in pink. Survey data were analyzed using Mann-Whitney. P < 0.05 is significantly different. *Abbreviations: F = funnel, B = bulb pipette*.

**Figure S5.**
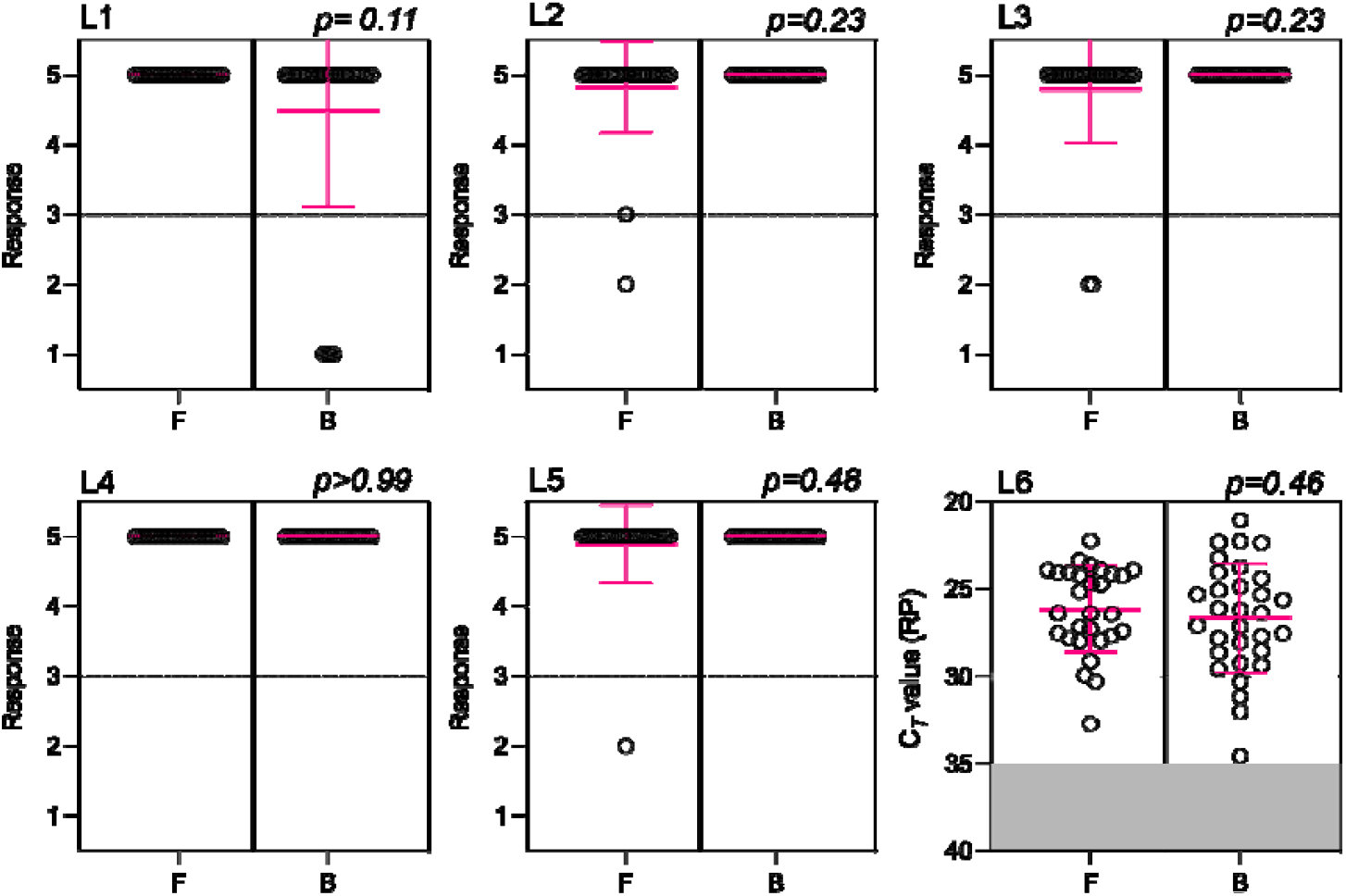
Responses to laboratory survey. Mean and standard deviation are marked in pink. P-values are shown for questions that could be assessed using Mann-Whitney. Ct values over 35 are considered invalid and are highlighted in gray on L6. *Abbreviations: F = funnel, B = bulb pipette*.

## Supplementary Tables

**Table S1.**
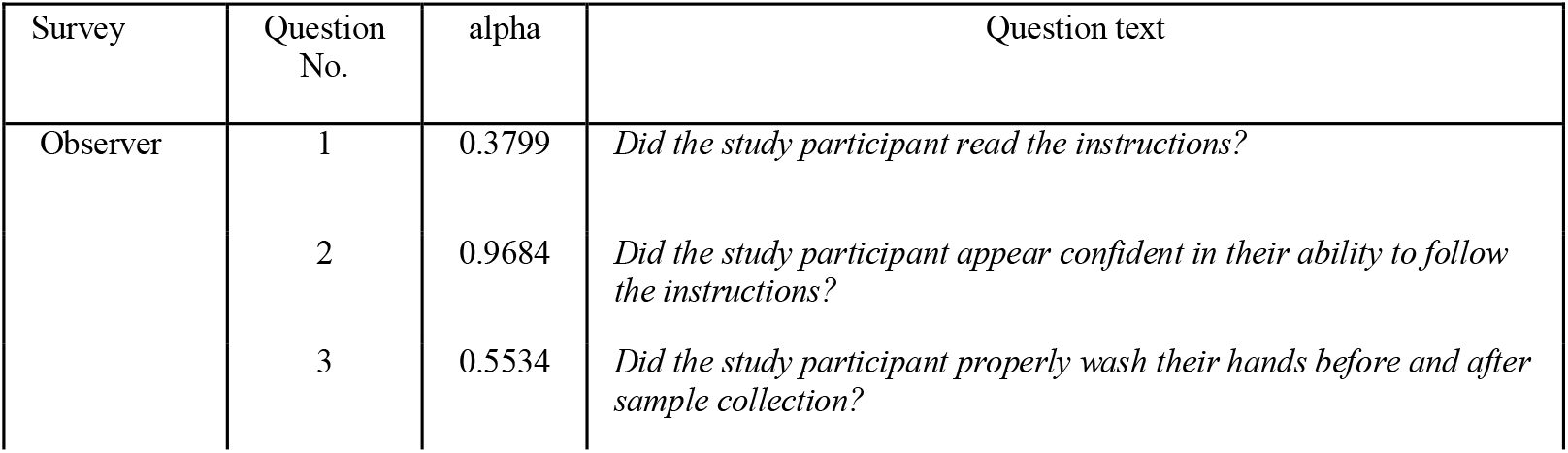
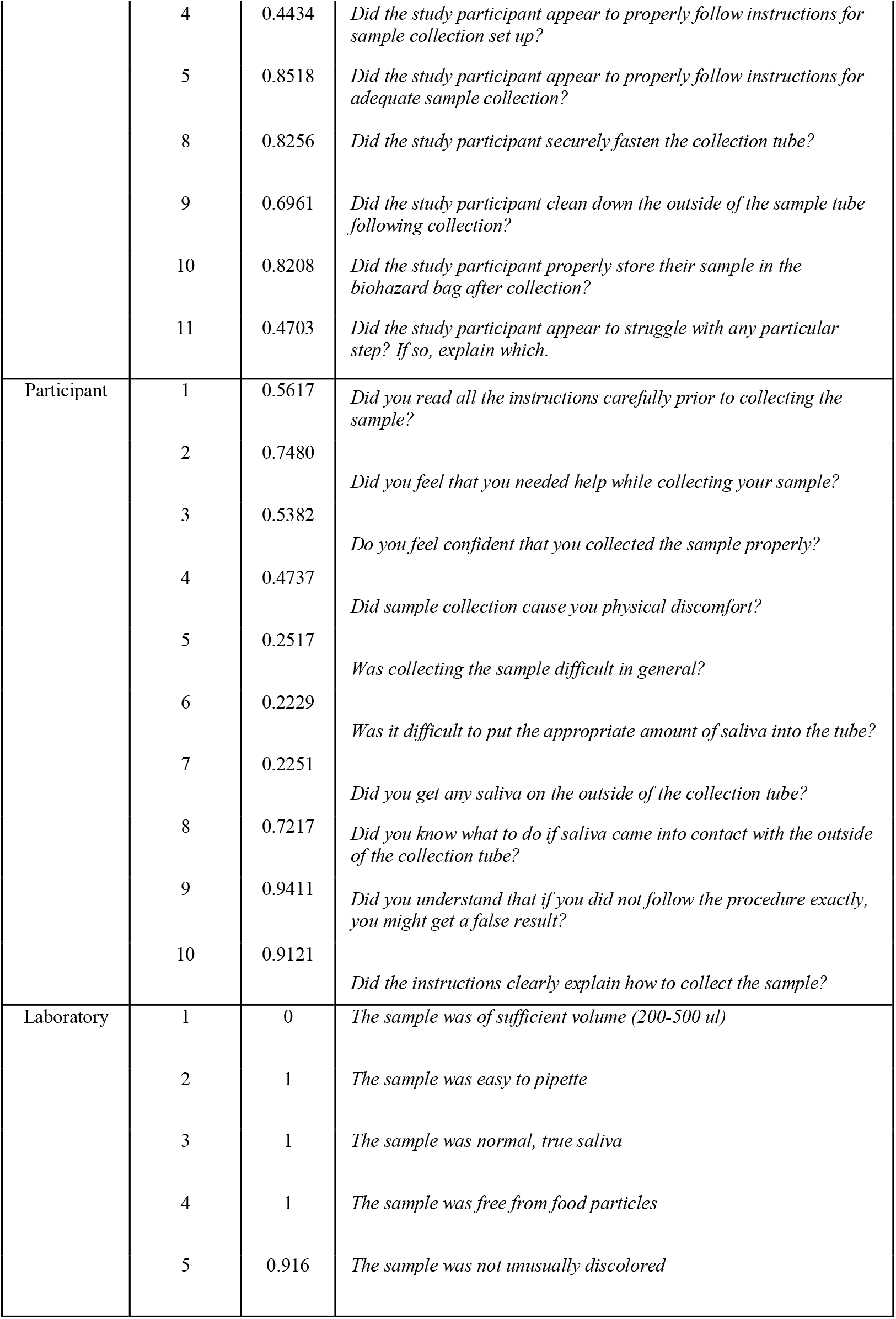
Internal reliability of survey question measured with Cronbach’s alpha.

**Table S2.** The majority of internally reliable survey questions did not differ significantly across collection devices. (a) Analysis of responses for participant and observer survey questions found to be internally reliable with Cronbach’s alpha. (b) Analysis of laboratory survey responses. P-values indicate the result of one-way ANOVA. Mean and standard deviation were reported for questions in which the response distribution was identical across devices. *Abbreviations: No. = number, st. dev. = standard deviation*

| a) | Survey | Question No. | P-value |
| --- | --- | --- | --- |
|  | Observer | 2 | 0.6668 |
|  |  | 3 | 0.2541 |
|  |  | 5* | 0.0267 |
|  |  | 8 | 0.2436 |
|  |  | 9 | 0.2256 |
|  |  | 10 | 0.3144 |
|  | Participant | 1 | 0.4736 |
|  |  | 2 | 0.7566 |
|  |  | 3 | 0.3315 |
|  |  | 8 | 0.5560 |
|  |  | 9 | 0.2612 |
|  |  | 10 | 0.2002 |

| b) | Question No. | Statistic | Value |
| --- | --- | --- | --- |
|  | 1 | P-value | 0.3256 |
|  | 2 | Mean (st. dev.) | 4.87 (0.346) |
|  | 3 | Mean (st. dev.) | 4.87 (0.346) |
|  | 4 | Mean (st. dev.) | 4.77 (0.568) |
|  | 5 | P-value | 0.4862 |
|  | 6 | P-value | 0.1192 |

**Table S3** Participant survey data for pilot study

**Table S4** Observation survey data for pilot study

**Table S5** Lab survey data for pilot study

**Table S6** Participant survey data for at home collection kit study

**Table S5** Lab survey data for at home collection kit study, including the ct values for the RNase P gene from the participant’s saliva samples

